# READi-Dem: ML-powered, web-interface tool for Robust, Efficient, Affordable Diagnosis of Dementia

**DOI:** 10.1101/2023.10.23.23297405

**Authors:** Verena Klar, Elinor Thompson, Melvin Selim Atay, Peter Owoade, Sofia Toniolo, Amir Dehsarvi, Sanjay Rathee

**Author notes:** Corresponding author: Sanjay Rathee. These authors contributed equally and designated as co-first authors (Verena Klar, Elinor Thompson). This author designated as project incharge (Sanjay Rathee).

## Abstract

**Background:** Dementia screening tools typically involve face-to-face cognitive testing. Indeed, this introduces an increasing burden on the clinical staff, particularly in low-resource settings. The objective of our study is to develop an integrated online platform for efficient dementia screening, using a brief and cost-effective assessment.

**Methods:** We used the Longitudinal Ageing Study in India dataset (LASI-DAD, n=2528) to predict dementia diagnosis based on the Clinical Dementia Rating (CDR). Using feature selection algorithms and principal component analysis (PCA), we identified key predictive features. We compared the performance of six machine learning (ML) classifiers that were trained on the 42 selected features (full model) and the two components identified by PCA (minimal model). The best-performing model was selected for our web platform.

**Results:** Selected features mapped onto two distinct, interpretable domains: a cognitive domain and an informant domain. The first two principal components cumulatively explained 90.2% of the variance and included questions from the Mini-Mental State Exam (MMSE) and the Informant Questionnaire on Cognitive Decline in the Elderly (IQCODE). Classifiers trained on the minimal model performed on par with the full model, with Support Vector Machine performing best (93.4%). The model did not reliably predict Parkinson’s disease (67% accuracy) or stroke (53.1% accuracy), suggesting dementia specificity. The respective questions from MMSE and IQCODE (27 items) were incorporated into our online platform.

**Conclusion:** We built an online platform enabling end-to-end screening for dementia from assessment to prediction, based on patient and caregiver reports. Web App code is available at GitHub: https://github.com/sanjaysinghrathi/READi-Dem & Web App link is available at Web Page: https://researchmind.co.uk/readi-dem. For the convenience of researchers, a video summarizing our work is also accessible on the Web App Page and [YouTube Link](https://youtu.be/jflT8Gm1xC8?si=AVudrAmi_4SOWTm1).

## 1 Introduction

DEMENTIA is a neurological syndrome that most commonly affects memory and cognitive functioning, leading to a progressive impairment of activities of daily living. Other symptoms include mood and personality changes, attention deficits, and language impairment.

In 2019, the prevalence of dementia in the older population was estimated to be 7.1% in the UK [1] and 9.8% in the US [2]. In low-resource settings, such as India, measuring dementia prevalence is more challenging, however, this rate is estimated to be up to 10.6% [3]. Due to ageing populations, the number of people living with dementia worldwide is predicted to rise from 57.4 million in 2019 to 152.8 million by 2050 [4].

As a result, the global financial burden of dementia is increasing: Global spending on dementia was estimated at 263 billion USD in 2019, attributable to diagnosis, treatment, and care costs [5]. This is predicted to further increase to 1.6 trillion USD by 2050, representing an estimated 11-17% of all healthcare spending [5]. In addition to the economic impact, the emotional and physical impact on patients, their caregivers, and family members is immense [6], [7].

Low-income countries are less well equipped to deal with the rising dementia burden than wealthy nations with strong healthcare provision. Dementia poses a particular challenge in India, due to its large population and extreme population growth. Furthermore, challenges in education lead to a low doctor-to-patient ratio, and environmental and lifestyle factors, such as pollution, lead to increased risk of dementia [3].

Early diagnosis is critical to promote early symptom management and appropriate triage to specialised support networks. Although there are currently no treatment options available to reverse the symptoms of dementia, it is likely that future treatments will need to be administered in the early stages of the disease before neurodegeneration advances [8]. Diagnostic processes can be highly heterogeneous across medical centers, and commonly involve face-to-face cognitive testing and expensive brain imaging. Currently, it is estimated that only one in ten people in India living with dementia receive a diagnosis or any specialised treatment [9], which highlights the difficulty of accessing suitable facilities in low-resource settings. Therefore, it is important to develop efficient and low cost systems for accurate diagnosis.

In this study, we leveraged machine learning (ML) to identify the minimum number of features needed to predict the outcome of a dementia screening, while avoiding those that are cost and labour intensive. We focused on the Longitudinal Aging Study in India - Diagnostic Assessment of Dementia (LASI-DAD) [10], [11]. The study follows the Harmonized Cognitive Assessment Protocol (HCAP) study design [12], designed by the US-based Health and Retirement Study. The dataset further incorporates comprehensive cognitive, informant, and general health measures [11], [13], providing a large set of features traditionally shown to be relevant for dementia diagnosis, as well as the clinical dementia rating (CDR), an assessment of dementia severity by clinicians.

A growing volume of research uses ML to predict diagnosis and progression of dementia from medical and clinical records [14], [15], [16]. One previous study has used the LASI-DAD dataset to explore ML methodologies for automatic dementia diagnosis. In this study, Jin et al. [17] performed several ML analyses for automatic diagnosis using the global CDR rating as the predictor, demonstrating the efficiency of support vector machines (SVMs) for dementia classification. However, they used a feature set containing summary scores from five different cognitive assessments, three assessments of self-reported functional difficulties, a depression and anxiety assessment, informant interview, socio-demographic variables, and the participants’ health history. As a result, their model cannot easily be transferred to a screening tool, as the data used in the model would be time-consuming and costly to collect, especially in a clinical setting.

In this study, we therefore sought to find a feature set with a minimal number of features that would allow for classification of dementia, while being economical to acquire. We first selected optimally predictive features. We further reduced the dimensionality of the dataset to only include the most meaningful components computed from the scores. Specifically, we sought to select items that rely neither on clinical interviews nor on cognitive testing, such that they could be performed by a caretaker. This ensures that our predictive models can be used as a time and cost-effective clinical screening tool.

Due to the harmonised nature of the HCAP, we were able to test the predictive utility of our feature set in an independent validation dataset the Aging, Demographics, and Memory Study (ADAMS) [18], acquired in the USA. This allowed us to explore the cross-cultural generalisation of the selected features and their applicability in a global context.

The field is gradually transitioning from face-to-face and time consuming cognitive assessments to digital screening tools, which are highly scalable and cost-effective. Hence, a large number of traditional screenings are available as computerised adaptations (see Thatbtah et al. for a review [19]). Here, as a proof-of-concept, we incorporated our best-performing ML model into a web platform. It provides a form to answer a set of questions that the model then uses predict a dementia diagnosis.

Our curated questionnaire and web platform enable rapid and affordable dementia diagnosis in the Indian population and our results offer preliminary evidence that this is robust across different global settings. Thus it can be used as a clinically useful computer aided diagnosis (CAD) tool.

## 2 Methods

A brief overview of the processing pipeline can be seen in Fig. 1.

**Fig. 1:**
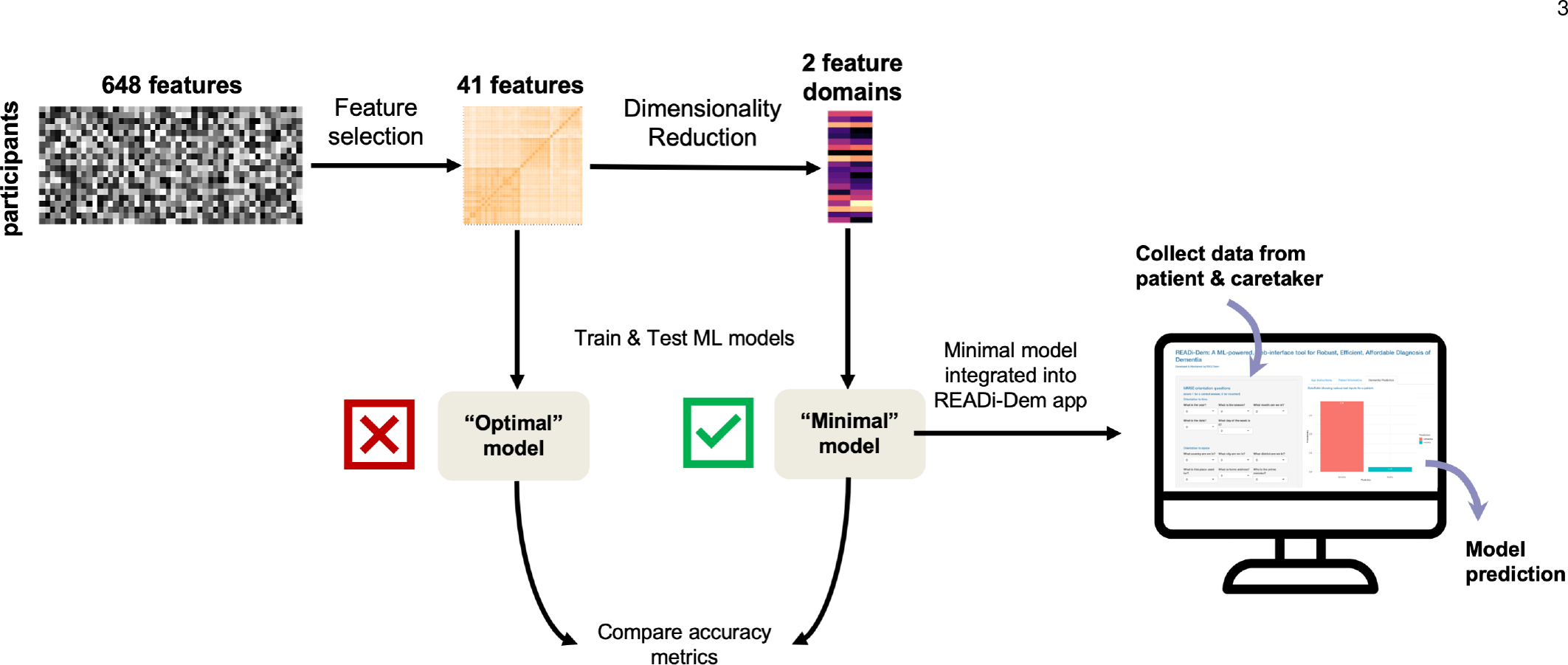
An overview of the analysis pipeline used in this paper. The Boruta feature selection algorithm was first used to select the best predictors for dementia as measured by the CDR, resulting in a reduced set of 41 features. In order to find the minimum number of necessary features, we applied PCA to reduce dimensionality further, identifying two distinct feature domains, cognitive functioning and activities of daily living, comprising 27 features. We trained and tested established ML models on the reduced (41 features) and the minimal feature sets (27) and compared performance metrics between them. The minimal model was incorporated into an online app: READI-DEM (Robust, Efficient, Affordable DIagnosis of DEMentia). This can be used to predict an individual’s likelihood of dementia diagnosis, using information that is quick and easy to obtain.

### 2.1 Data

#### 2.1.1 LASI-DAD

We used data from the Longitudinal Ageing Study in India Diagnostic Assessment of Dementia (LASI-DAD) [10], [11], [13]. A total of N = 2528 participants took part in the study (age *M* = 69.7, *SD* = 7.6), living in 18 out of 28 Indian states. 60% of the cohort lived in rural areas and the majority did not complete secondary education. In order to create a dataset with a sufficient number of dementia cases, those at risk of dementia were oversampled. The dataset comprised 730 variables in total. In broad categories, this included measures of cognitive health (e.g., Hindi Mental State Exam), Informant measures of dementia and daily living (e.g., Blessed Dementia Scale), and general Geriatric Assessment (weight and height). A full breakdown of assessments can be found in Table 5.

Based on the available measures, a dementia diagnosis was given by three independent neurologists according to the Clinical Dementia Rating (CDR) [20]. The final score for each patient was agreed upon by all three clinicians or by a moderator in case of disagreement. The CDR rates the participants’ impairment as 0 – None, 0.5 – Questionable, 1 – Mild, 2 – Moderate, 3 – Severe, made up by 30.40%, 62.03%, 6.41%, 0.99%, 0.20% of the sample, respectively (Fig. 2).

**Fig. 2:**
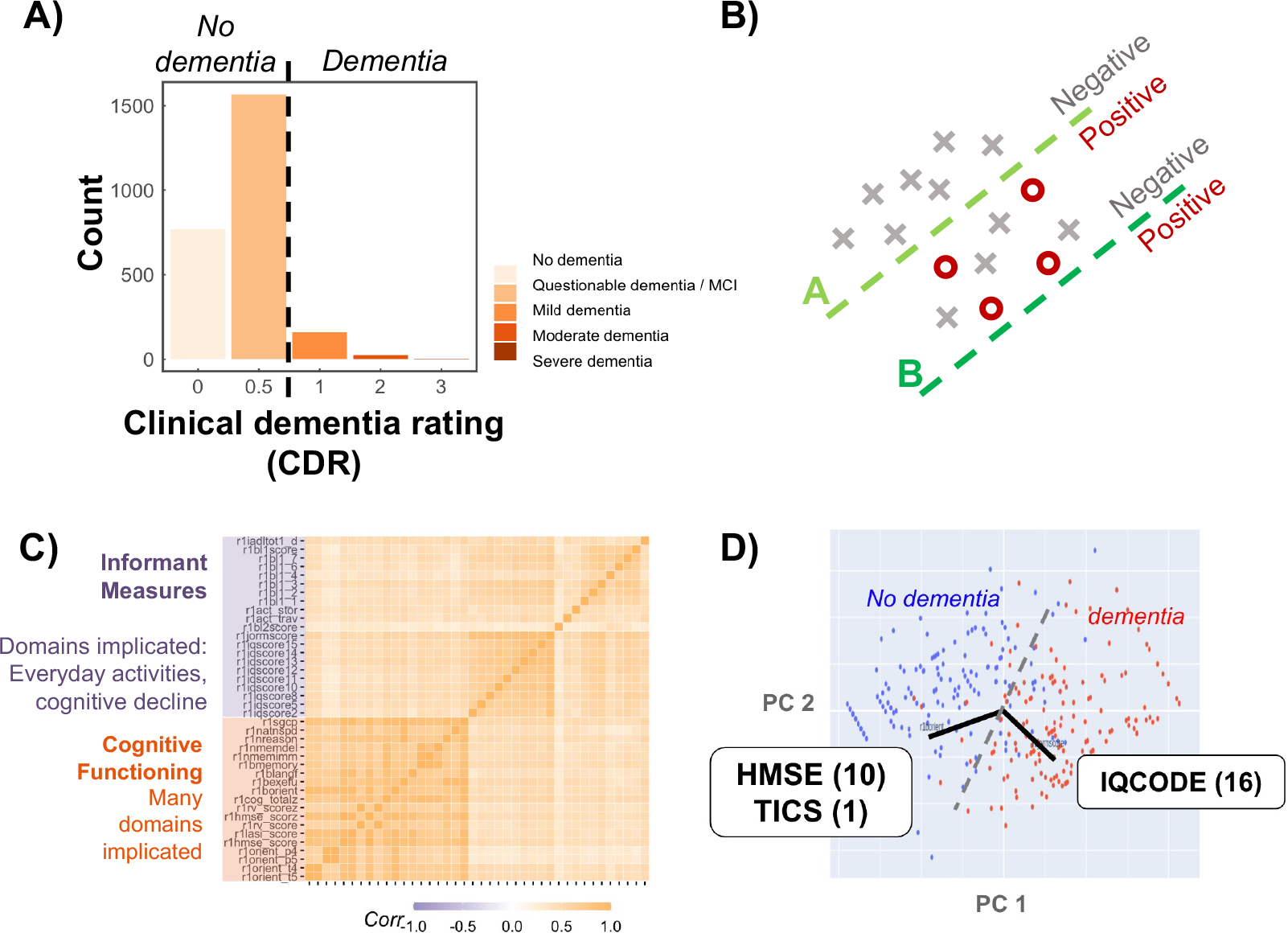
A) Prevalence of dementia in the LASI-DAD sample according to the Clinical Dementia Rating (CDR). The dotted line represents our binary criterion. B) The effect of class imbalance on separation line. Without accounting for class imbalance, classifiers would choose a cut-off favouring prediction of “negative” (non-dementia, line B) instead of achieving a meaningful prediction (line A). C) Correlation between features selected by the Boruta algorithm1. The features map onto to distinct clusters: measures of cognitive functioning (orange) and measures of everyday activities (purple). D) Principal-component analysis showed that the space described by the top two principal components was equally well spanned by a combination of only 27 items from the Hindi Mental Status Exam (HMSE), Telephone Interview for Cognitive Status (TICS), and the Informant Questionnaire on Cognitive Decline in the Elderly (IQCODE). These items were selected to collect data from patients and caregivers on our final platform, in the form of a few simple questions (figure 1).

#### 2.1.2 ADAMS

As a validation dataset, we used the Aging, Demographics, and Memory Study (ADAMS) [18], collected in the USA and linked to the nationally representative Health and Retirement Study (HRS). We used data from N = 854 participants aged 70 or above, who received an extensive in-home clinical and neuropsychological assessment to determine their dementia status. We performed manual feature matching to find the corresponding features in the ADAMS dataset to those we had selected for the minimal feature set in the LASI-DAD. The model was consequently trained on the ADAMS dataset and tested on the set aside validation set.

### 2.2 Pre-processing

#### 2.2.1 Missing Data

We filtered out features with more than 25% missing values, so as not to introduce unnecessary bias. Missing values for the remaining features were imputed using the median. In order to implement a binary predictive model of dementia, we binarized the sample into dementia (*CDR* ≥ 1) and non-dementia (*CDR <* 1) as shown in Fig. 2A. After pre-processing, the resulting dataset contained 704 variables.

#### 2.2.2 Class Imbalance

Imbalanced classes refer to a disproportionate ratio of observations in each class and are one of the common problems in biological datasets. Fig. 2B shows an example where class imbalance can cause a false cutoff point. Most ML models aim to minimise the classification error for a given cutoff point. If class imbalance is not addressed, the optimal cutoff is the one that always predicts the outcome to be the over-represented class. The LASI-DAD cohort is highly imbalanced with 192 dementia patients and 2336 non-dementia samples (Fig. 2A). To address this, we used down-scaling to balance the power between dementia and non-dementia samples – analysing data from 172 subjects in each category at a time and rotating through examples in the non-dementia group. The down-scaled dataset was used for the feature selection and model selection steps.

### 2.3 Feature Selection

Our dataset involves 648 features that could be used as disease markers to predict dementia. However, many of them are either insignificant, interrelated, or codependent, and they could be a source of unwanted noise during ML model training. Therefore, we used Boruta, a random forest-based R package (v7.0.0) for feature selection. This technique is most beneficial with ML models that are lacking stringent feature selection capabilities [21]. The Boruta feature selector extracted 41 most significant features (out of 648 features) for the classification of subjects according to their dementia diagnosis. A total number of 237 features are required to derive this “optimal feature set”, as some of these are summary scores derived from multiple features.

#### Algorithm 1 Pre-Processing & Feature Selection

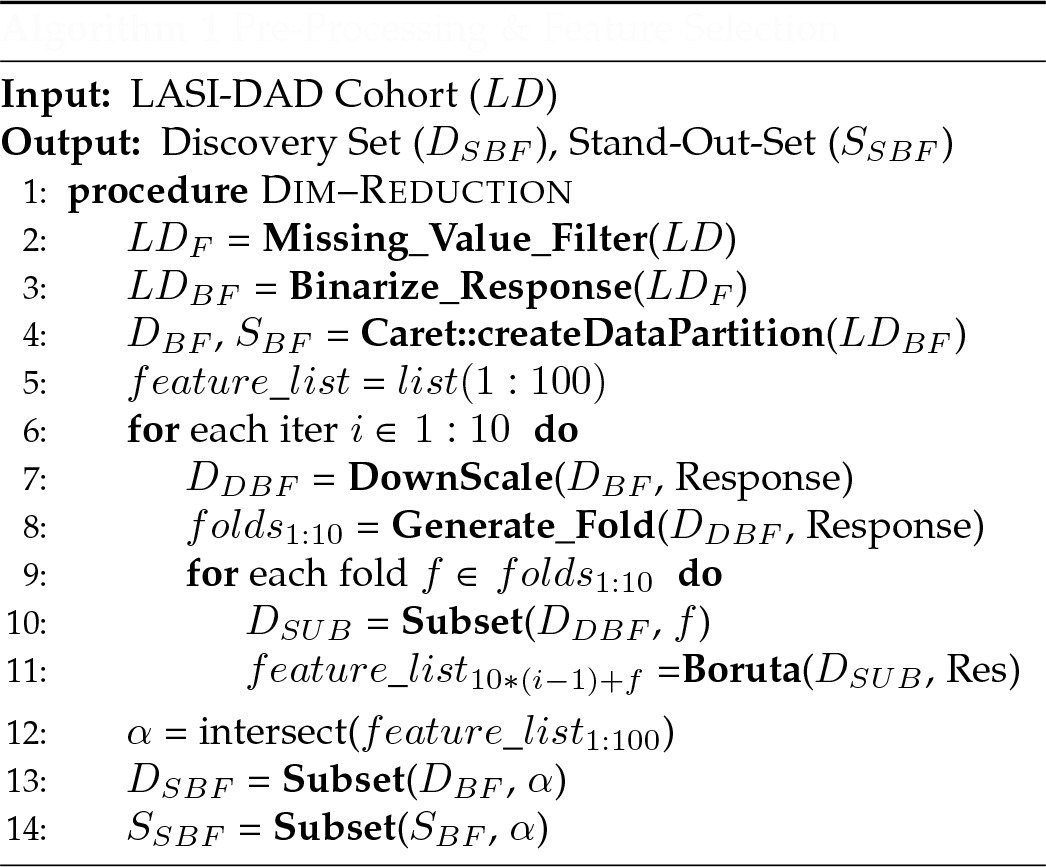

The algorithm 1 highlights the pseudo code for preprocessing and feature selection steps. The analysis starts with filtering missing data and binarising the response column for LASI-DAD data (lines 2-3). The pre-processed cohort is then partitioned as a discovery set (*D*_*BF*_) and a stand-out-set (*S*_*BF*_) with 80:20 ratio (line 4). A list of size 100 (line 5) is initialised to store the significant features for two iterative loops, each of length 10. The first loop handles the class imbalance (lines 6-7) without losing any data power by selecting all the 194 dementia samples, but it uses different 194 non-dementia samples in each iteration. The second loop handles the outlier effect (lines 9-10) by selecting a random selection of 172 (80%) samples from the demented and non-demented samples. The Boruta algorithm is executed 100 times over different sets of 344 samples to find the most predictive features (line 11). At last, the features that are found significant in all the iterations are used to create new discovery and stand-out sets, upon having selected the features column only with binarised responses and without any missing data (lines 13-14).

### 2.4 Feature Refinement

The features selected by the Boruta algorithm were highly correlated and formed two clusters (Fig. 2C), which neatly mapped onto two domains: the cognitive functioning and the activities of daily life. Given our aim of reducing the time and monetary cost needed to administer the screening, we further reduced the features using PCA. Based on the outcome, we retained the first two components, respectively explaining 57.2% and 20.2% of the variance (77.4% variance explained cumulatively).

Incidentally, the new 2-dimensional space was well spanned by two compound features from our 41 Boruta-selected features (Fig. 2D): (1) A compound feature of the first 10 items from the Hindi Mental State Exam (HMSE [22]) and 1 item from the Telephone Interview for Cognitive Status (TICS [23]); (2) The compound score of the Informant Questionnaire on Cognitive Decline in the Elderly, computed from 16 items (IQCODE [24]); comprising 27 items in total. Consequently, these measures are easy to acquire in the form of simple questions without the need to perform any interviews or cognitive tests for the patients. These items are listed in Table 1.

**TABLE 1:** Questionnaire items included in the minimal feature set. Items included from the Hindi Mini Mental State Exam (HMSE), Telephone Interview for Cognitive Status (TICS), and the Informant Questionnaire on Cognitive Decline in the Elderly (IQCODE).

| Questionnaire | Item number | Item code: LASI-DAD | Item code: ADAMS | Description | ADAMS description (if different) |
| --- | --- | --- | --- | --- | --- |
| HMSE | 1 | RwYR | ANMSE1 | what is the year? |  |
| HMSE | 2 | RwSEASON | ANMSE2 | what is the season? |  |
| HMSE | 3 | RwDATE | ANMSE3 | what is the date? |  |
| HMSE | 4 | RwDW | ANMSE4 | what is the day of the week? |  |
| HMSE | 5 | RwMO | ANMSE5 | what month is it? |  |
| HMSE | 6 | RwSTATE | ANMSE6 | what state are we in? |  |
| HMSE | 7 | RwCITY | ANMSE7 | what city are we in? | what county are we in? |
| HMSE | 8 | RwFLOOR | ANMSE8 | what floor of the building are we on/ what is this place used for? | what city are we in? |
| HMSE | 9 | RwADDRESS | ANMSE9 | what is your home address? | What hospital are we in? |
| HMSE | 10 | RwNAME | ANMSE10 | what is the name of your district? | What floor are we on? |
| TICS | 3 | RwPRIME | ANPRES | what is the name of the current prime minister? | Who is the President of the United States right now? |
| IQCODE | 1 | R1IQSCORE1 | AGQ14 | ability to remember things about family and friends |  |
| IQCODE | 2 | R1IQSCORE2 | AGQ15 | ability to remember things that have happened recently |  |
| IQCODE | 3 | R1IQSCORE3 | AGQ16 | ability to recall conversations a few days later |  |
| IQCODE | 4 | R1IQSCORE4 | AGQ17 | ability to remember their address and telephone number |  |
| IQCODE | 5 | R1IQSCORE5 | AGQ18 | ability to remember what day and month it is |  |
| IQCODE | 6 | R1IQSCORE6 | AGQ19 | ability to remember where things are usually kept |  |
| IQCODE | 7 | R1IQSCORE7 | AGQ20 | ability to remember where to find things that have been put in a different place from usual |  |
| IQCODE | 8 | R1IQSCORE8 | AGQ21 | ability to know how to work familiar machines around the house |  |
| IQCODE | 9 | R1IQSCORE9 | AGQ22 | ability to learn to use a new gadget or machine around the house |  |
| IQCODE | 10 | R1IQSCORE10 | AGQ23 | ability to learn new things |  |
| IQCODE | 11 | R1IQSCORE11 | AGQ24 | ability to follow a story in a book or on TV |  |
| IQCODE | 12 | R1IQSCORE12 | AGQ25 | ability to make decisions on everyday matters |  |
| IQCODE | 13 | R1IQSCORE13 | AGQ26 | ability to handle money for shopping |  |
| IQCODE | 14 | R1IQSCORE14 | AGQ27 | ability to handle financial matters |  |
| IQCODE | 15 | R1IQSCORE15 | AGQ28 | ability to handle everyday arithmetic problems |  |
| IQCODE | 16 | R1IQSCORE16 | AGQ29 | ability to use intelligence to understand what's going on and to reason things through |  |

### 2.5 Machine Learning (ML) Models

In order to test their predictive accuracy, we used the 27 refined features to predict the binarised CDR. In other words, a total number of 27 features (representing the whole dataset) and one binary dementia class for 344 samples (balanced by dementia class) was thus selected as the discovery cohort. This cohort was then divided as training (80%) and stand-out sets (20%). The training set with 276 samples was consequently used to identify the best ML model for our data.

Finally, we trained different ML algorithms package: logistic regression, ridge regression, lasso regression, elastic net, support vector machines (SVMs), neural networks, random forest, gradient boosting machines (GBM), and k-nearest neighbors (KNN). The models were tested on our discovery cohort and evaluated based on k-fold cross-validation. We validated the model with the highest accuracy rate and precision on the stand-out set.

The algorithm 2 details pseudo code used to calculate prediction accuracy rate for different ML models. The cohort for this step is taken from the output of algorithm 1. The procedure starts with generating a data frame to store iteration number, the cross-validation accuracy for all six ML models, and the same six ML model accuracy rates on stand-out-set (line 2). It repeats the same iterative loop approach as in algorithm 1 to handle the class imbalance and the outlier effects (lines 3-6). It runs multiple ML models on training data (instead of running Boruta) to find the k-fold cross validation and stand-out-set accuracy rates (lines 7-15). At last, the average of the cross validation accuracy rates is calculated to identify the best model (line 19).

### 2.6 RShiny Online Platform

Our online platform was built in RShiny [25] and is hosted at researchmind.co.uk/readi-dem. Features from the best performing model can be collected ad hoc via a form. The responses are then fed into the model and a prediction is returned and visualised in the form of a barplot showing the likelihood that the participant has dementia.

#### Algorithm 2 ML Model Training

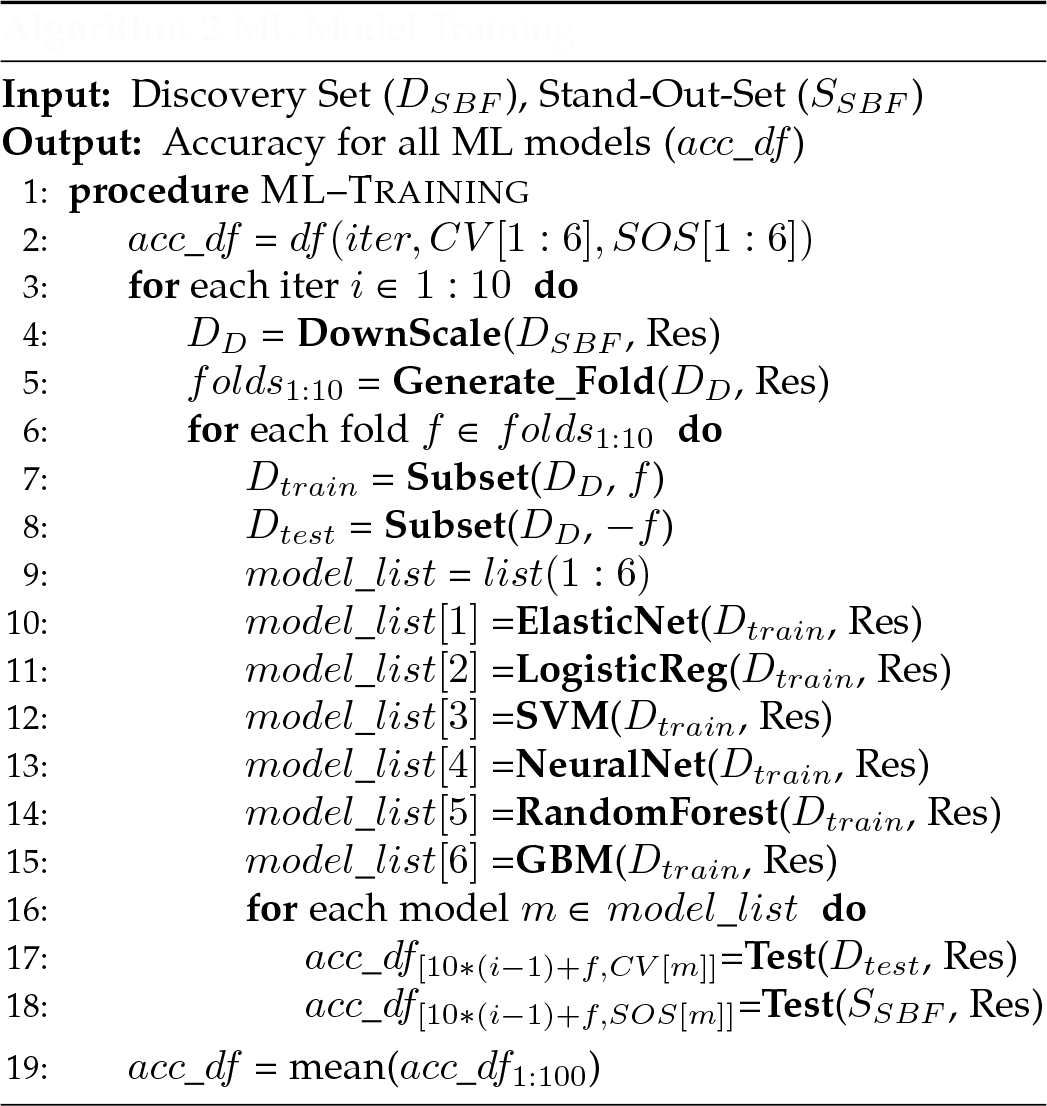

## 3 Results

### 3.1 Minimal feature set yields competitive accuracy rates despite stark reduction in features

We calculated the cross-validation predictive accuracy of different ML models on the down-scaled dataset, using the optimal feature set (41 features) and the minimal feature set (27 features). The optimal feature set contained compound scores that would take 237 individual items (e.g. questionnaire items and cognitive test scores) to compute. Results are shown in (Table 2). Crucially, models trained on the minimal feature set performed on par with the ones trained on the optimal feature set, with the best model reaching a stand-out set accuracy of 95.6% in both cases. In terms of different ML models, all performed well, with cross validation accuracy rates exceeding 87%. We found that the SVMs (89.7%) yielded the highest classification accuracy rate in the cross validation analysis. Furthermore, upon testing on the stand-out dataset, SVMs had a predictive accuracy of 94.1% when using the minimal feature set.

**TABLE 2:** Classification accuracy rates for different ML models on the minimal feature set with balanced classes.

| Dataset | Model | Testing Method | elastic | logistic | svm loov | neuralnet | random forest | gradient boosting |
| --- | --- | --- | --- | --- | --- | --- | --- | --- |
| LASI | Optimal | cross-validation | 0.934 | 0.860 | **0.937** | 0.926 | 0.911 | 0.913 |
| LASI | Optimal | stand-out set | **0.956** | 0.868 | **0.956** | **0.956** | **0.956** | **0.956** |
| LASI | Minimal | cross-validation | 0.894 | 0.896 | **0.897** | 0.895 | 0.868 | 0.876 |
| LASI | Minimal | stand-out set | 0.941 | **0.956** | 0.941 | **0.956** | 0.911 | 0.941 |

**TABLE 3:** Performance metrics of the support vector machine model on the stand-out dataset, comparing between optimal and minimal feature sets. The optimal feature set comprises 41 selected features that were computed from 237 items. This minimal feature set contains 27 refined features.

| Feature set | Sensitivity | Specificity | Precision | Recall | F1 Score |
| --- | --- | --- | --- | --- | --- |
| Optimal | 0.94 | 0.97 | 0.97 | 0.94 | 0.96 |
| Minimal | 0.97 | 0.79 | 0.83 | 0.97 | 0.89 |

**TABLE 4:** Classification accuracy rates for different ML models with the minimal feature set in the ADAMS dataset.

| Dataset | Testing Method | elastic | logistic | svm loov | neuralnet | random forest | gradient boosting |
| --- | --- | --- | --- | --- | --- | --- | --- |
| ADAMS | cross-validation | 0.842 | 0.827 | 0.807 | 0.874 | **0.910** | 0.908 |
| ADAMS | stand-out set | 0.837 | 0.815 | 0.837 | 0.880 | 0.880 | **0.902** |

### 3.2 Predictive power is dementia-specific rather than disease-general

In order to rule out our models distinguishing between healthy and diseased rather than predicting dementia specifically, we tested the models on two control groups. We used the informant-provided information in the LASI-DAD dataset to determine which individuals had been diagnosed with stroke (n = 164) and Parkinson’s disease (n = 112). We then performed the same cross validation accuracy tests using the trained model as described above, with the diagnosis results for stroke and Parkinson’s as the labels for the model. The model differentiated the Parkinson’s patients and stroke patients from healthy controls with an accuracy rate of 67% and 53.1%, respectively. This suggests that the predictive power of our model is specific to dementia rather than disease-general.

### 3.3 Feature selection for minimal models generalises to external dataset

We retrained our models on a different dataset to find out if the same features were predictive of dementia in an independent dataset. In order to achieve this, we matched our minimal feature set to features in the the Aging, Demographics, and Memory Study (ADAMS) [18]. The corresponding features are shown in Table 1. With the minimal feature set, we were able to achieve a cross validation accuracy of 91% in the ADAMS dataset, using the random forest algorithm.

### 3.4 App User Interface provides predictions for new individuals

We built an online platform that uses our minimal machine learning model to predict the likelihood of a dementia diagnosis for an individual, based on patient and caregiver reports. The app can be accessed via https://researchmind.co.uk/readi-dem. The user interface (UI) was designed to be clear and easy to use with simple instructions. It consists of 11 questions from the MMSE and 16 from the IQCODE included in our minimal model. The responses to these questions are used as inputs for the machine learning model, and consequently, a probability for dementia diagnosis is displayed. A representative user interface is shown in figures 1, 3 and 4. The scripts for the analyses and the platform are all available at https://github.com/sanjaysinghrathi/READi-Dem.

## 4 Discussion

In this study, we developed a web-based application to aid in low-cost dementia screening, using established ML techniques of the unique population-level Indian LASI-DAD dataset. We identified the most strongly-predictive features for dementia diagnosis from our dataset, and further refined them to a minimal set of 27 features using dimensionality reduction, placing emphasis on selecting question-based measures. We evaluated the predictive power of a variety of ML approaches with a support-vector machine model reaching the highest binary classification accuracy in a cross validation analysis. Crucially, our selected minimal feature set was on par with using larger feature sets. Predictive power was not replicated in Parkinson’s and stroke control groups, suggesting dementia-specific prediction. The chosen features were validated in a similar, US-based ADAMS dataset, with models reaching well above chance accuracy rates. The best-performing model trained on the minimal feature set was incorporated into an online platform on which users can answer a standard questionnaire to predict the likelihood of a dementia diagnosis for in a new individual. The study thus provides big-data validation of the value of simple, established questionnaires in a developing and a developed country. The screening process does not require cognitive testing and provides a fast and accurate prediction of dementia outcome, making it especially suitable for use in low-income countries.

Dementia screening platforms and digital apps that utilise machine learning have shown promising results in detecting early signs of cognitive decline and dementia and thus identifying individuals at risk of dementia. Our study suggests that it only takes 27 items to achieve an accuracy comparable with more extensive testing. This is a large improvement upon previous studies. For example, a similar study conducted on the LASI-DAD dataset used the full feature set available (29 features comprised of summary scores from 127 different items) [17]. Their models reached reached a similar accuracy to ours at 95%. This is despite the fact that our optimal and minimal feature sets being a 3-fold and 5-fold reduction in the number of items that need to be collected compared to the full set, respectively. This finding highlights the importance of feature selection in the development of tools for clinical contexts, where time is scarce, and suggests that the combination of IQCODE and MMSE items we found may be most efficient given those constraints.

Further, a recent Korean study [16] followed a similar path to ours. They developed an ML algorithm that is able to identify both mild cognitive impairment and dementia based on neuropsychological screening test results, specifically the MMSE, Montreal Cognitive Assessment (MoCA), Korea Dementia Screening Questionnaire (KDSQ). A combination of these screenings reached extremely high accuracy rates distinguishing controls from MCI and dementia. A drawback of this is that in a clinical setting, despite overlap in the questions, all three screenings would have to be used, all of them involving, albeit short, cognitive tests that require a clinician present. Our study shows that similar performance can be achieved with a screening consisting only of verbal questions that might not need a clinician.

Recently, there as been growing interest in harmonising datasets concerned with ageing and dementia [26]. This is a difficult undertaking as even when studies follow the Harmonized Cognitive Assessment Protocol (HCAP), features found in one data set may be missing from the other. Despite this, cross-validation accuracy rates on this validation dataset reached 91%. This suggests features may generalise cross-culturally even between culturally dissimilar nations. It also suggests that optimising screenings while keeping them to a minimal length may ultimately improve translatablity. Many of our selected features have a cross-national range of application, with the MMSE and IQCODE having been validated in a multitude of countries [27], [28], [29], [30]. Optimising feature sets is therefore crucial to minimise not only the cost of application, but the cost of validation of screening tools in low-resource settings.

The web platform developed in this study is capable of classifying dementia patients within seconds. It is easy to use and can be accessed using any web browser of choice on a computer or a smartphone. Screening for neurodegenerative diseases can be time consuming and requires a trained specialist. To address this timely issue, our platform can be easily used by a caregiver regardless of their clinical training level. Since it is online, assessment can happen remotely, for example in the patient’s home. This is especially beneficial for countries with difficult health access due to lack of resources, infrastructure, high population, or geographical size. Although the current language is English, the possibility of translation is promising, as discussed in the section on cross-cultural generalisation.

There are several limitations to the study and the resulting online tool. Firstly, in light of extant assessments, the question might arise why another assessment is relevant. For example, the general Practitioner Assessment of Cognition (GPCOG) [31] uses a mixture of informant and patient questions just like ours, and has been implemented in a website [32]. However, in contrast to our assessment, it also includes short cognitive testing elements, such as the clock drawing test. In addition, our questionnaire further validates these measures through its data-driven approach, promising robustness.

Secondly, there are some drawbacks to our choice of outcome measure. Since we used the CDR, diagnoses in the study were based on clinical decisions, with no imaging confirmation. As a result, our measure predicts clinical opinions rather than ground-truth dementia state. While we found that the questionnaire was specific to detecting dementia specificity by applying it to Parkinson’s disease and stroke, it cannot be used to distinguish different forms of dementia, such as Alzheimer’s, vascular, or Lewy-body dementia. Moreover, the rating was binarized, our tool is also not suited for the detection of mild cognitive impairment (MCI) which is often a precursor for dementia. James et al. [15] demonstrated successful machine learning models (SVMs) in medical settings using National Association of Care Catering (NACC) prognostic study data, but their model struggles with sensitivity (47%). Future research could explore the use of feature reduction in predicting dementia progression and its integration into online tools.

Thirdly, it is of note that the current version of the app is a proof of concept and further work is required to complete the design and functionality of the app, as well as address ethical and privacy concerns. Furthermore, a major limitation with such platforms is a potential lack of standardisation in the screening process such as different settings and test-givers.

## 5 Conclusion

We presented a minimal set of 27 questionnaire items comprised of patient and informant measures, which provides predictions of dementia diagnosis comparable with more extensive screenings. Formal cognitive testing is not required, allowing administration by a caregiver, such as a family member. Results can be made available within a few seconds via a web app. Future research could extend our approach to enable efficient screening for MCI, distinction of dementia sub-types, and prediction of disease progression. Moreover, this study contributes to generalising dementia screenings and improving their implementation to different countries, in particular developing countries with lack of healthcare access.

## Data Availability

All data used for ML model are available online at https://lasi-dad.org

## 6 Acknowledgements

We would like to acknowledge the DEMON Network for their kind and continuous support with this research from forming the idea in the DEMON NeuroHack 2022 until the dissemination of this research. Our special thanks goes to the University of Exeter for the Alan Turing grant to fund this project. We would like to express our appreciation to the Milner Therapeutics Institute at the University of Cambridge for their invaluable support in overseeing the financial management of the grant.

### APPENDIX

**Fig. 3:**
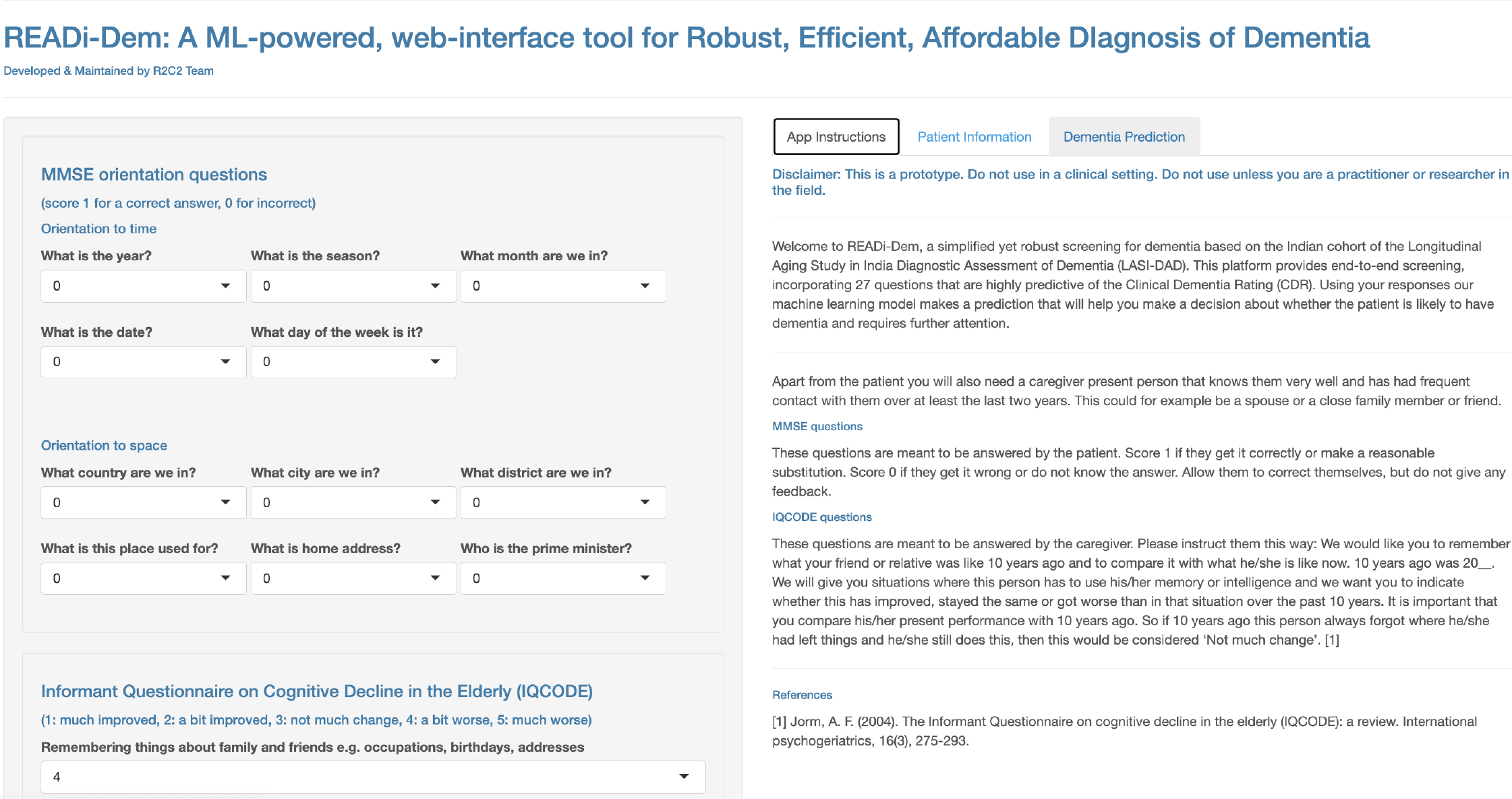
A sample image of the user interface of the platform. The caregiver can easily enter the information in the required fields in order to generate a final report.

**TABLE 5:**
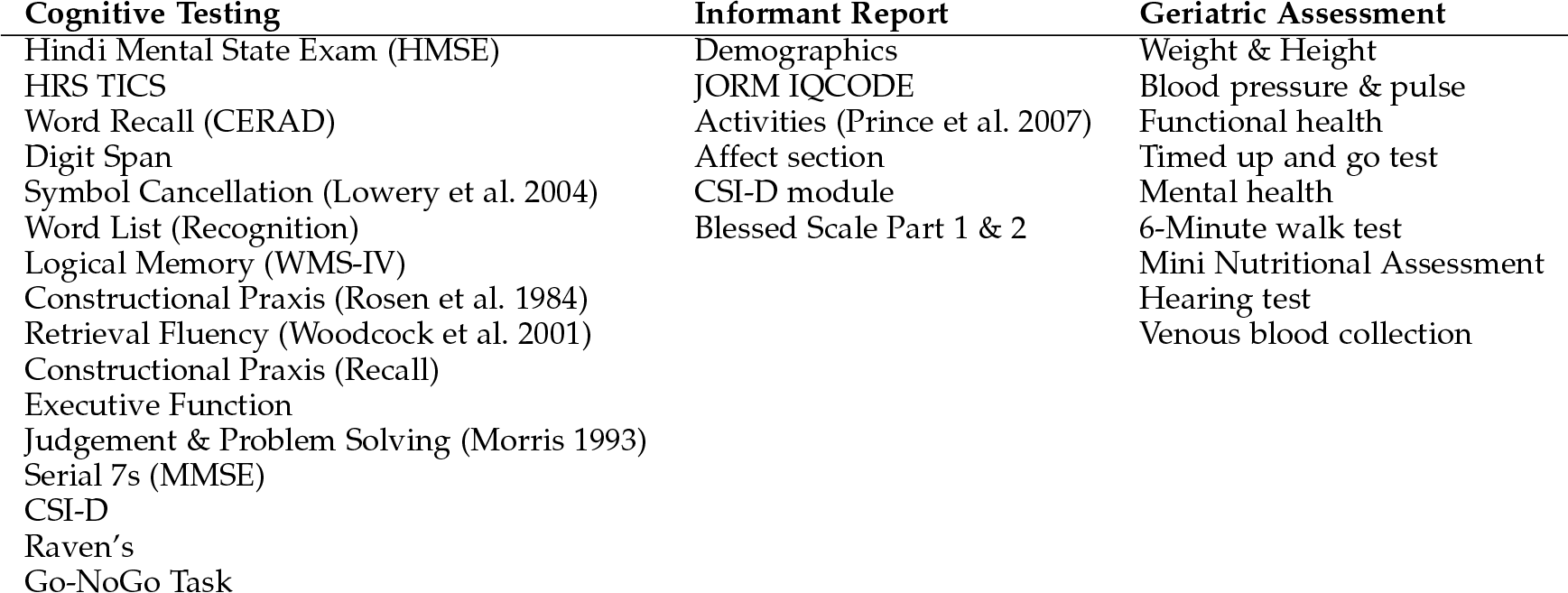
Domains tested as part of the LASI-DAD study.

| Cognitive Testing | Informant Report | Geriatric Assessment |
| --- | --- | --- |
| Hindi Mental State Exam (HMSE) | Demographics | Weight & Height |
| HRS TICS | JORM IQCODE | Blood pressure & pulse |
| Word Recall (CERAD) | Activities (Prince et al. 2007) | Functional health |
| Digit Span | Affect section | Timed up and go test |
| Symbol Cancellation (Lowery et al. 2004) | CSI-D module | Mental health |
| Word List (Recognition) | Blessed Scale Part 1 & 2 | 6-Minute walk test |
| Logical Memory (WMS-IV) |  | Mini Nutritional Assessment |
| Constructional Praxis (Rosen et al. 1984) |  | Hearing test |
| Retrieval Fluency (Woodcock et al. 2001) |  | Venous blood collection |
| Constructional Praxis (Recall) |  |  |
| Executive Function |  |  |
| Judgement & Problem Solving (Morris 1993) |  |  |
| Serial 7s (MMSE) |  |  |
| CSI-D |  |  |
| Raven's |  |  |
| Go-NoGo Task |  |  |

**TABLE 6:**
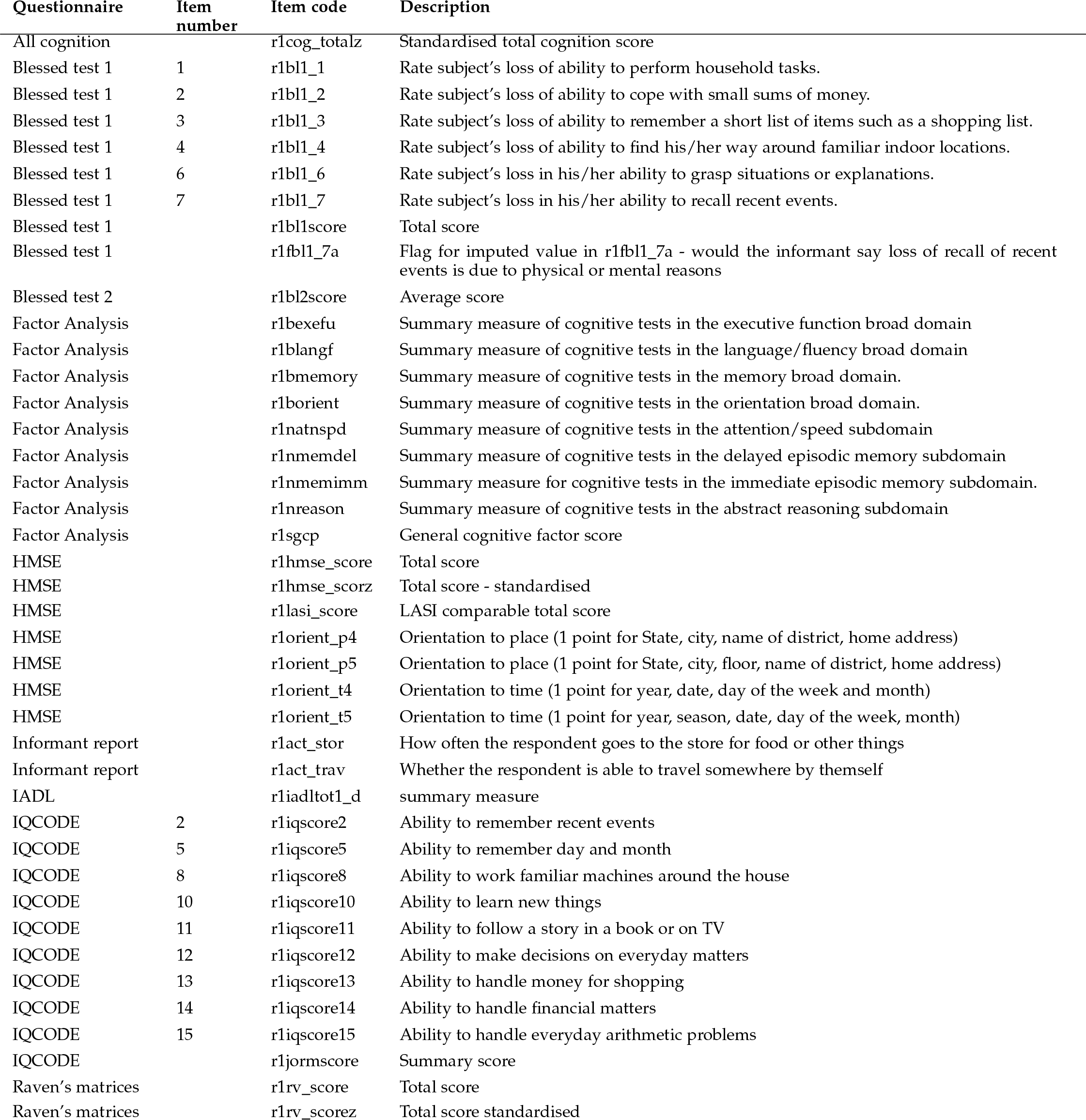
Questionnaire items included in the optimal feature set, identified with the Boruta algorithm from the LASI-DAD dataset. Items are included from: the Blessed test; a factor analysis of cognitive tests; the Hindi Mental State Examination (HMSE); an informant report on everyday activities; the Instrumental Activities of Daily Living (IADL); the Informant Questionnaire on Cognitive Decline in the Eldery (IQCODE); and the Raven’s matrices test.

**Fig. 4:**
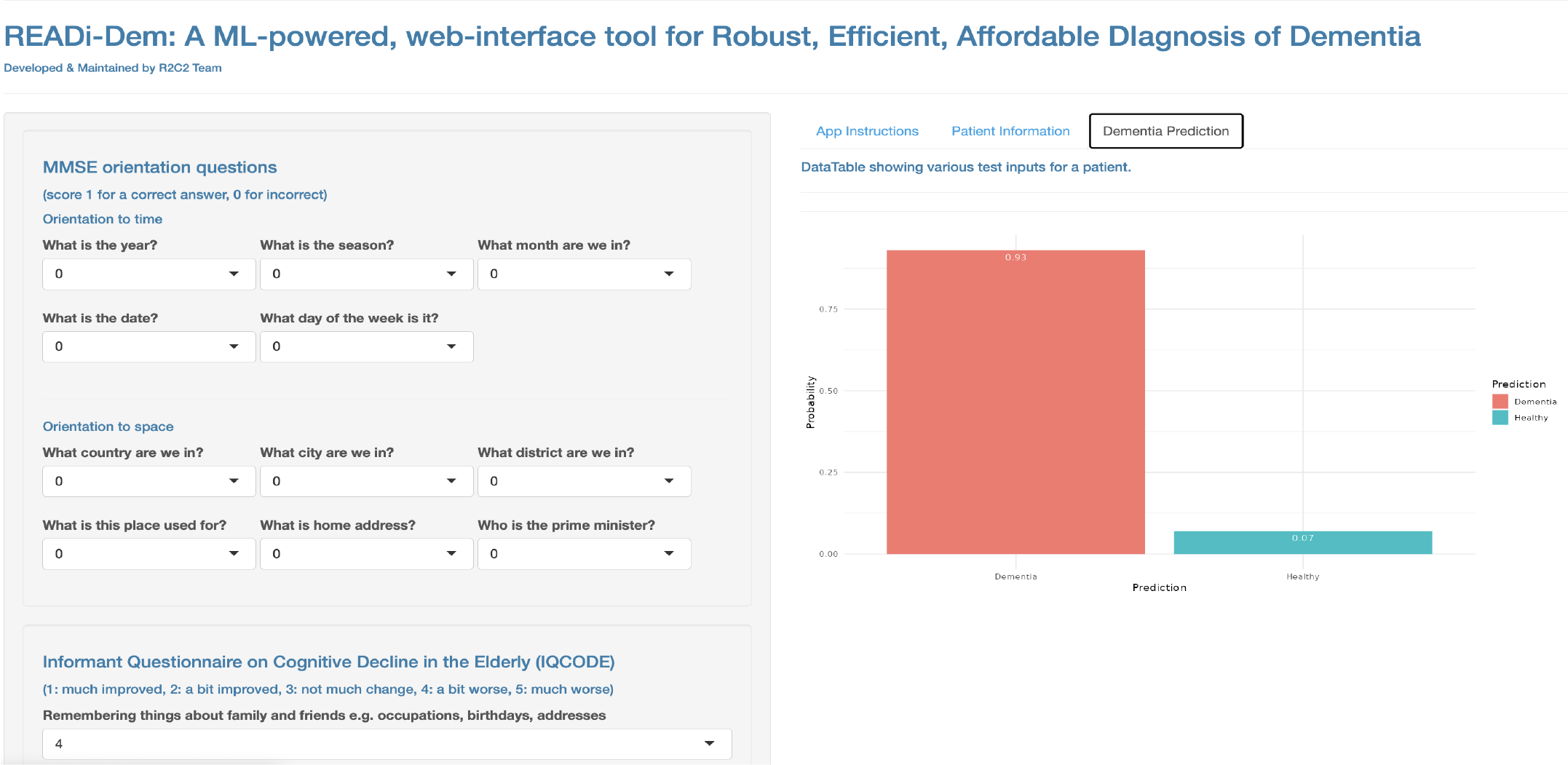
A sample image of the user interface of the platform for dementia prediction.

**TABLE 7:** Items contained in the final questionnaire included in the online platform. HMSE: Hindi Mental State Exam, TICS: Telephone Interview of Cognitive Status, IQCODE: Informant Questionnaire on Cognitive Decline.

|  | Source | Item |
| --- | --- | --- |
| 1 | HMSE | Is it morning or afternoon or evening? |
| 2 | HMSE | What day of the week is it today? |
| 3 | HMSE | What date is it today? |
| 4 | HMSE | Which month is it today? |
| 5 | HMSE | What season of the year is it? |
| 6 | HMSE | Under which post office does your village come? |
| 7 | HMSE | Which district does your village fall under? |
| 8 | HMSE | Which village are you from? |
| 9 | HMSE | Which block (If village has any blocks) OR Which numbered area is this? |
| 10 | HMSE | Which place is this? |
| 11 | TICS | Who is the current prime minister? |
| 12 | IQCODE | Remembering things about family and friends, e.g. occupations, birthdays, addresses |
| 13 | IQCODE | Remembering things that have happened recently |
| 14 | IQCODE | Recalling conversations a few days later |
| 15 | IQCODE | Remembering her/his address and telephone number |
| 16 | IQCODE | Remembering what day and month it is |
| 17 | IQCODE | Remembering where things are usually kept |
| 18 | IQCODE | Remembering where to find things which have been put in a different place from usual |
| 19 | IQCODE | Knowing how to work familiar machines around the house |
| 20 | IQCODE | Learning to use a new gadget or machine around the house |
| 21 | IQCODE | Learning new things in general |
| 22 | IQCODE | Following a story in a book or on TV |
| 23 | IQCODE | Making decisions on everyday matters |
| 24 | IQCODE | Handling money for shopping |
| 25 | IQCODE | Handling financial matters, e.g. the pension, dealing with the bank |
| 26 | IQCODE | Handling other everyday arithmetic problems, e.g. knowing how much food to buy, knowing how long between visits from family or friends |
| 27 | IQCODE | Using his/her intelligence to understand what's going on and to reason things through |

**TABLE 8:** Performance metrics of the 6 ML models comparing between optimal and minimal feature sets. Optimal performance is based on the set of 41 selected features that were computed from 237 items. Minimal performance is based on the set of 27 refined features.

| Dataset | Model | Testing Method | elastic | logistic | svm loov | neuralnet | random for-est | gradient boosting |
| --- | --- | --- | --- | --- | --- | --- | --- | --- |
| LASI | Optimal | sensitivity/specificity | 0.94/0.97 | 0.82/0.91 | **0.94/0.97** | 0.94/0.97 | 0.94/0.97 | 0.94/0.97 |
| LASI | Optimal | precision/recall/F1 Score | 0.97/0.94/0.96 | 0.90/0.82/0.86 | **0.97/0.94/0.96** | 0.97/0.94/0.96 | 0.97/0.94/0.96 | 0.97/0.94/0.96 |
| LASI | Minimal | sensitivity/specificity | 0.97/0.82 | 0.97/0.82 | **0.97/0.79** | 0.97/0.79 | 0.97/0.82 | 0.94/0.82 |
| LASI | Minimal | precision/recall/F1 Score | 0.85/0.97/0.90 | 0.85/0.97/0.90 | **0.83/0.97/0.89** | 0.83/0.97/0.89 | 0.85/0.97/0.90 | 0.84/0.94/0.89 |

